# Waning Effectiveness of SARS-CoV-2 mRNA Vaccines in Older Adults: A Rapid Review

**DOI:** 10.1101/2022.01.15.22269364

**Authors:** Etsuro Nanishi, Ofer Levy, Al Ozonoff

## Abstract

The U.S. Centers for Disease Control and Prevention (CDC) and other health agencies have recently recommended a booster dose of COVID-19 vaccines for specific vulnerable groups including adults 65 years and older. There is limited evidence whether vaccine effectiveness in older adults decreases over time, especially against severe COVID-19. We performed a rapid review of published studies available through 04 November 2021 that provide effectiveness data on mRNA vaccines approved/licensed in the United States and identified eight eligible studies which evaluated vaccine effectiveness in older adults. There is evidence of a decline in vaccine effectiveness against both SARS-CoV-2 infection and severe COVID-19 in older adults among studies which analyzed data up to July-October 2021. Our findings suggest that vaccine effectiveness diminishes in older adults, which supports the current recommendation for a booster dose in this population.

## Introduction

Authorized/licensed COVID-19 mRNA vaccines had shown remarkable efficacy and real-world effectiveness against both asymptomatic and symptomatic disease caused by severe acute respiratory syndrome coronavirus 2 (SARS-CoV-2) including in adults aged 65 or older ^1, 2^. However, accumulating evidence suggests that vaccine effectiveness (VE) against SARS-CoV-2 infection declines over time in some groups, including older adults who have the highest risk of morbidity and mortality from COVID-19 ^3^. This waning effectiveness, along with the emergence of variants of concern (VOC), may be associated with a resurgence of COVID-19 cases ^4, 5^. On 22 September 2021, the FDA granted Emergency Use Authorization (EUA) for booster doses of the Pfizer BioNtech mRNA vaccine in specific populations including older adults 65 years and older; long-term care setting residents; those who have high risk of severe COVID-19; and those who work or live in high-risk settings ^6, 7^. On 20 October 2021 a similar EUA was granted regarding Moderna boosters. There are relatively few published studies regarding the durability of SARS-CoV-2 mRNA VE in older adults with respect to protection against severe COVID-19 leading to hospitalization and death.

Several studies have reported VE of older adults against SARS-CoV-2 infection and severe COVID-19. Each study varies in terms of vaccines, geographical regions or countries, and observation period after immunization. Each of these factors may potentially affect generalizability of findings to other settings. Furthermore, dominant circulating strains of SARS-CoV-2 during the study period may vary between studies. To the best of our knowledge, there is no comprehensive review to assess waning VE in older adults, representing a knowledge gap with important implications for vaccination policy. We therefore reviewed and synthesized available data of VE among older adults for currently approved/licensed mRNA vaccines in the USA (i.e. BNT162b2 and mRNA1283) from peer-reviewed published PubMed-indexed studies that were available through 04 November 2021. Aiming to enhance the evidence base informing public health policy, we assessed VE in adults stratified by study period and with an emphasis on the older age group.

## Methods

We conducted a PubMed search on 04 November 2021 for English-language articles, using combinations of the search terms of “SARS-CoV-2”, “COVID-19”, “vaccine”, “effectiveness”, “efficacy”, “older adults”, and “elder”. We also searched reference lists of identified articles and other relevant articles on effectiveness of COVID-19 vaccines in older adults. We included human studies which evaluated VE of the two mRNA vaccines employed in the U.S. (BNT162b2 and mRNA1273) in adults aged 65 or older. Given considerable variation between countries in key factors such as timing of vaccine campaigns, modification of vaccine regimens (e.g. prolonged interval between immunizations), and circulating SARS-CoV-2 strains, we limited our review to U.S.-based studies. We excluded preprint papers and studies which only report VE after single dose, and only considered estimates of VE at least 7 days after second vaccination.

We captured three key elements from each eligible study: 1) definition and estimate of VE; 2) observation period; and 3) number of participants. We also captured VE estimates for younger adults if included in the same article. We classified each study definition of VE as protection from any SARS-CoV-2 infection versus protection from severe COVID-19 defined as hospital admission or death. We defined three calendar periods based in part on the emergence of the Delta variant of SARS-CoV-2: 1) the ‘early’ period before the Delta variant was identified (14 December 2020 to 05 May 2021); ‘middle’ period as Delta became more widespread (14 December 2021 to 05 August 2021); and the ‘late’ period during which Delta was the predominant circulating virus (01 July 2021 to 31 October 2021).

## Results

After screening titles, abstracts, and full-text reviews, we identified a final set of N=8 published articles that met our eligibility criteria. All articles evaluated VE of adults ≥65 years of age, while six of eight (88%) studies also analyzed VE in younger adults. One study evaluated VE for BNT162b2 vaccine, while six evaluated VE for BNT162b2 and mRNA1273 vaccines either separately or combined. All but one paper was classified as ‘early’ or ‘middle’ period studies (Table 1).

**Table 1.**
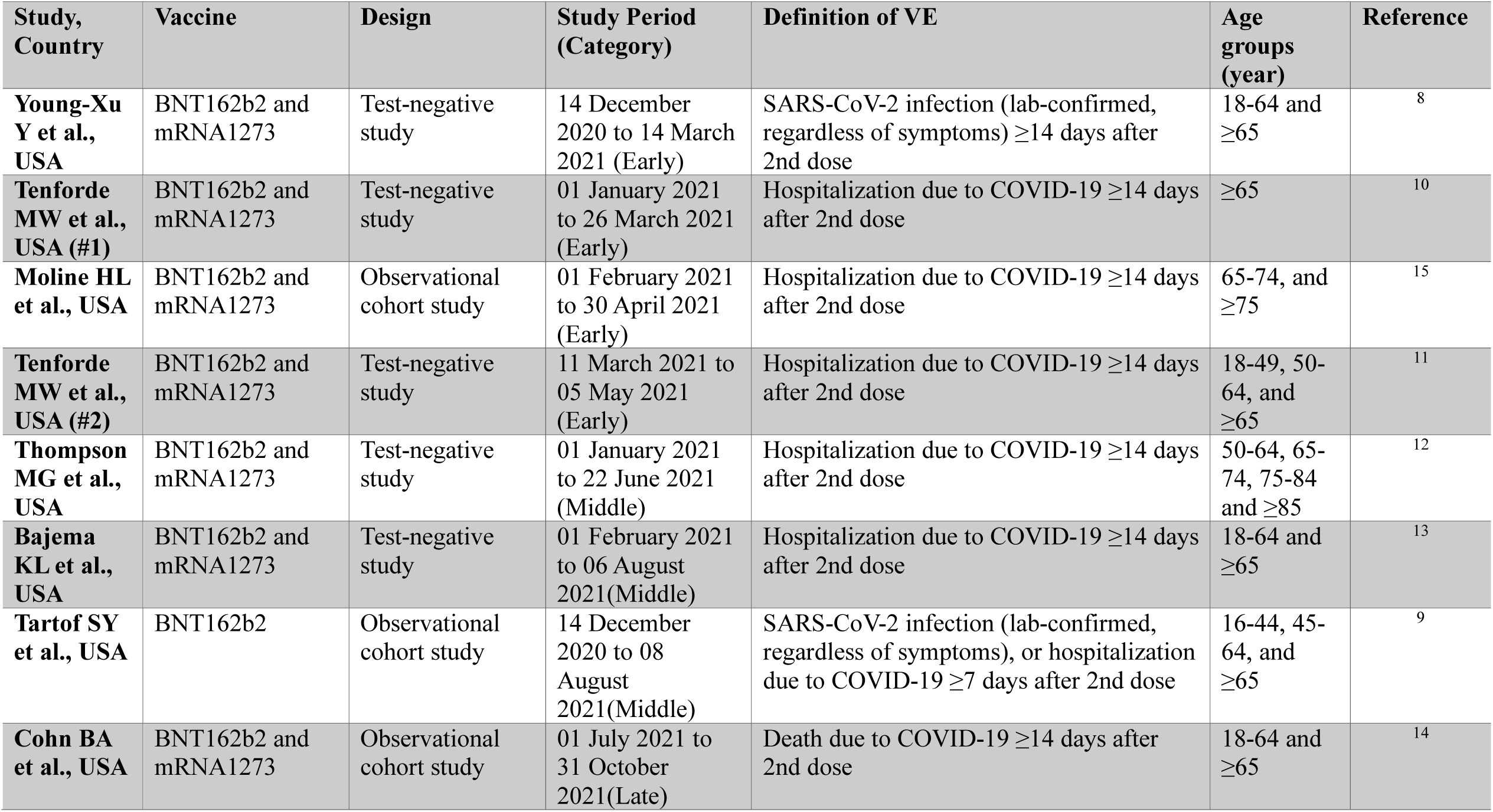
Study characteristics.

| **Study, Country** | **Vaccine** | **Design** | **Study Period (Category)** | **Definition of VE** | **Age groups (year)** | **Reference** |
| --- | --- | --- | --- | --- | --- | --- |
| **Young-Xu Y et al., USA** | BNT162b2 and mRNA1273 | Test-negative study | 14 December 2020 to 14 March 2021 (Early) | SARS-CoV-2 infection (lab-confirmed, regardless of symptoms) $\geq 14$ days after 2nd dose | 18-64 and $\geq 65$ | <sup>8</sup> |
| **Tenforde MW et al., USA (#1)** | BNT162b2 and mRNA1273 | Test-negative study | 01 January 2021 to 26 March 2021 (Early) | Hospitalization due to COVID-19 $\geq 14$ days after 2nd dose | $\geq 65$ | <sup>10</sup> |
| **Moline HL et al., USA** | BNT162b2 and mRNA1273 | Observational cohort study | 01 February 2021 to 30 April 2021 (Early) | Hospitalization due to COVID-19 $\geq 14$ days after 2nd dose | 65-74, and $\geq 75$ | <sup>15</sup> |
| **Tenforde MW et al., USA (#2)** | BNT162b2 and mRNA1273 | Test-negative study | 11 March 2021 to 05 May 2021 (Early) | Hospitalization due to COVID-19 $\geq 14$ days after 2nd dose | 18-49, 50-64, and $\geq 65$ | <sup>11</sup> |
| **Thompson MG et al., USA** | BNT162b2 and mRNA1273 | Test-negative study | 01 January 2021 to 22 June 2021 (Middle) | Hospitalization due to COVID-19 $\geq 14$ days after 2nd dose | 50-64, 65-74, 75-84 and $\geq 85$ | <sup>12</sup> |
| **Bajema KL et al., USA** | BNT162b2 and mRNA1273 | Test-negative study | 01 February 2021 to 06 August 2021 (Middle) | Hospitalization due to COVID-19 $\geq 14$ days after 2nd dose | 18-64 and $\geq 65$ | <sup>13</sup> |
| **Tartof SY et al., USA** | BNT162b2 | Observational cohort study | 14 December 2020 to 08 August 2021 (Middle) | SARS-CoV-2 infection (lab-confirmed, regardless of symptoms), or hospitalization due to COVID-19 $\geq 7$ days after 2nd dose | 16-44, 45-64, and $\geq 65$ | <sup>9</sup> |
| **Cohn BA et al., USA** | BNT162b2 and mRNA1273 | Observational cohort study | 01 July 2021 to 31 October 2021 (Late) | Death due to COVID-19 $\geq 14$ days after 2nd dose | 18-64 and $\geq 65$ | <sup>14</sup> |

### VE against SARS-Cov-2 Infection among Older Adults

Two studies examined mRNA VE against SARS-CoV-2 infection among older adults (Table 1) ^8,9^. One early study evaluated VE between 14 December 2020 and 14 March 2021, reporting that mRNA vaccines demonstrated VE of 93% among older adults against SARS-CoV-2 infection (Figure 1) ^8^. In contrast, a middle period study which evaluated VE during 14 December 2020 and 08 August 2021 showed considerable decline of VE against SARS-CoV-2 infection to 61% (Figure 1) ^9^.

**Figure 1.**
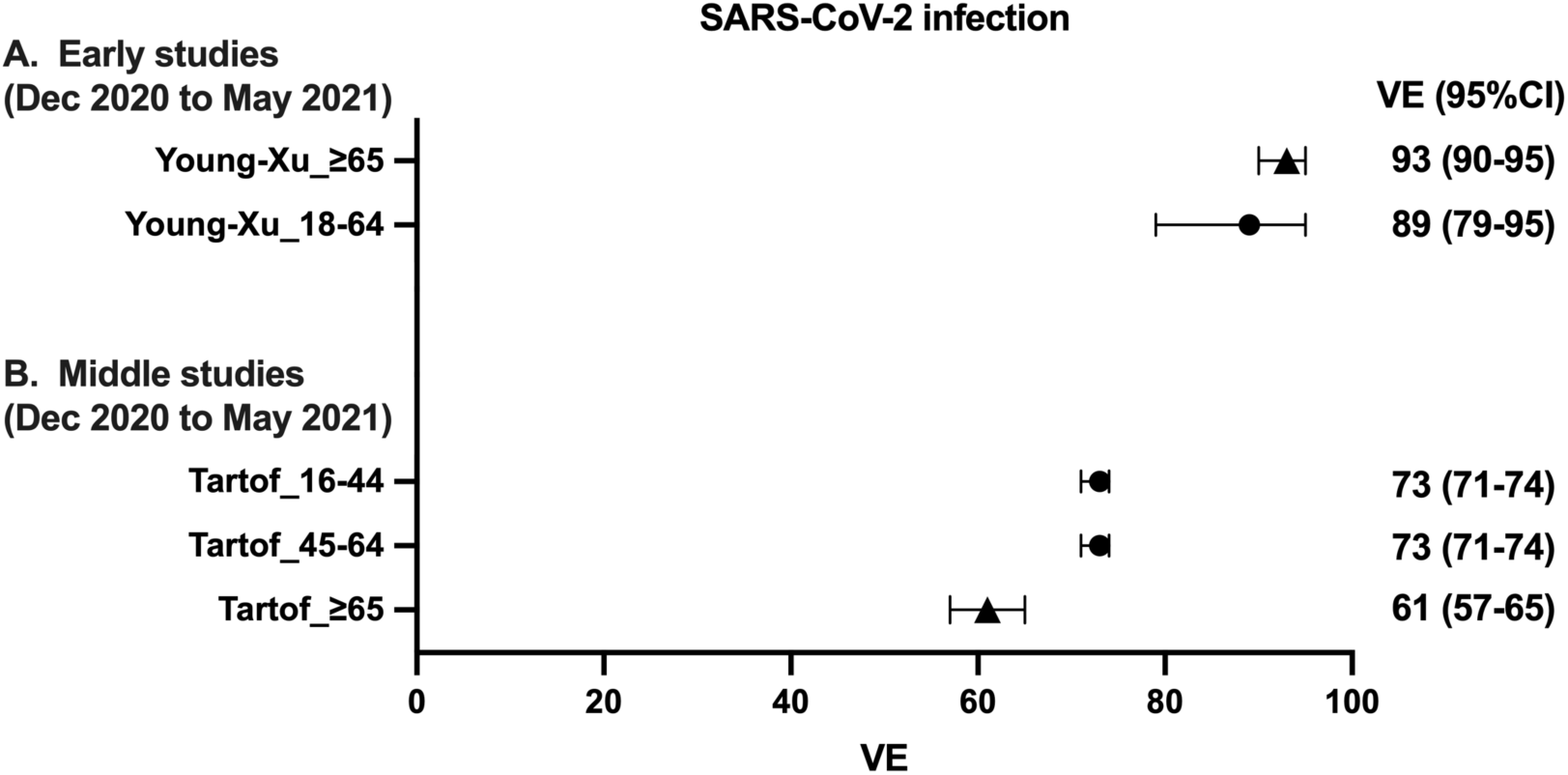
Forest plots of the vaccine effectiveness against SARS-CoV-2 infection. Point estimates and confidence intervals are depicted for mRNA vaccine effectiveness (VE) against SARS-CoV-2 infection. VE for BNT162b2 and mRNA1273 were evaluated at least 7 days after the second vaccination. Studies performed from 14 Dec 2020 up to 05 May 2021 and 08 Aug 2021 were categorized as “early” and “middle” studies, respectively. Circle symbols represents data for young adults (<65 years of age) and triangle symbols represents data for older adults (≥65 years of age). VE: vaccine effectiveness, CI: confidence interval.

### VE against Severe COVID-19 among Older Adults

Seven studies examined mRNA VE against severe COVID-19 among older adults (Table 1) ^9-15^. Among the three early studies, estimates of VE against severe COVID-19 ranged from 87.3% to 96.8% ^10, 11, 15^. Three middle studies evaluated VE up to 08 August 2021 (Table 1). Estimates of VE for middle studies ranged from 80% to 90% among older adults (Figure 2) ^9, 12, 13^. One study evaluated VE of mRNA vaccines among US veterans during 01 July to 31 October 2021, and was categorized as late study ^14^. The study evaluated VE against severe COVID-19 defined as death and reported marked decline in VE for older adults ranging from 70.1% (for BNT162b2) to 75.5% (for mRNA1273) (Figure 2).

**Figure 2.**
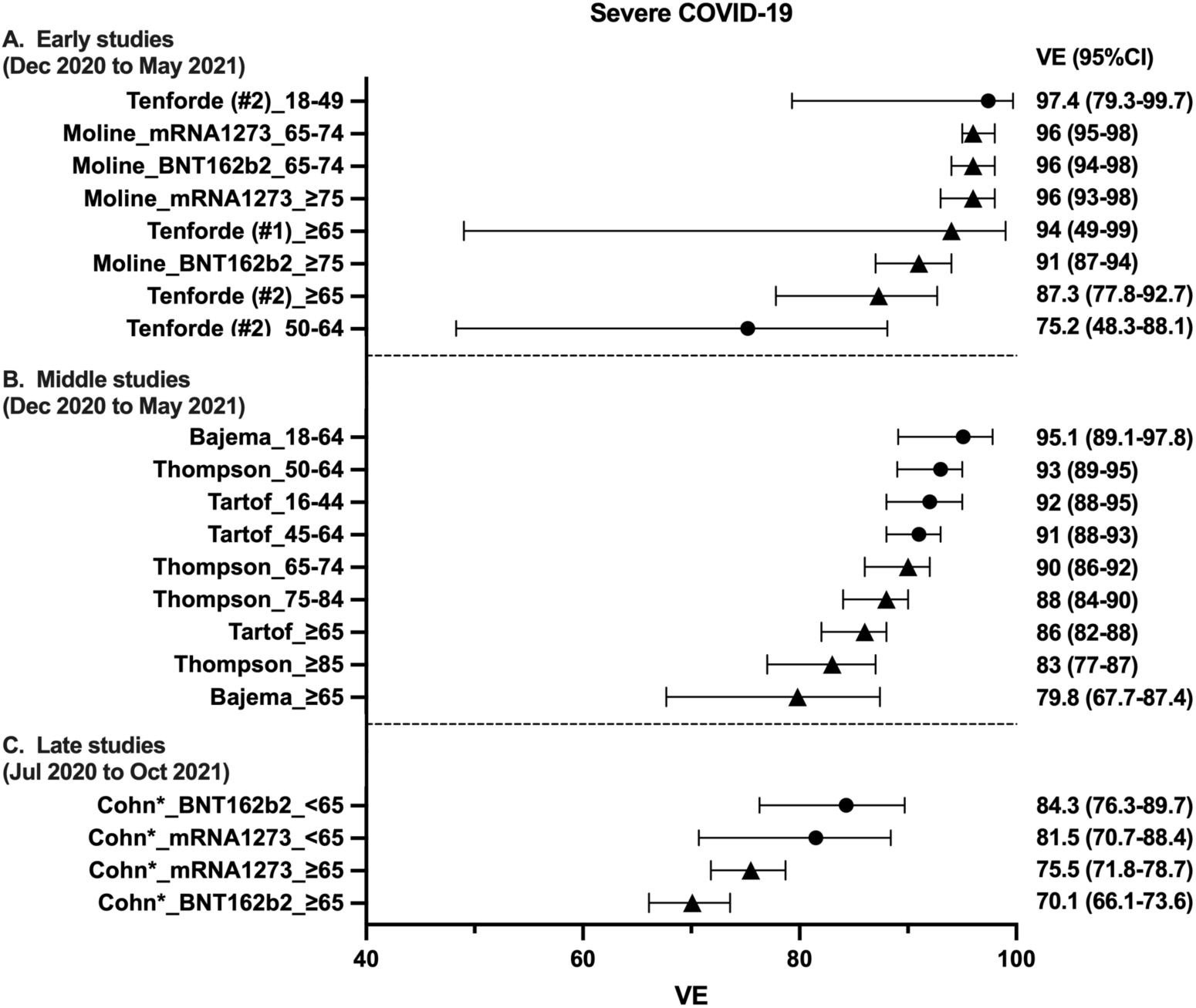
Forest plots of mRNA vaccine effectiveness against severe COVID-19. Vaccine effectiveness (VE) data against severe COVID-19 are shown. BNT162b2 and mRNA1273 mRNA VE were evaluated at least 7 days after the second vaccination. Studies performed from 14 Dec 2020 up to 05 May 2021 and 08 Aug 2021 were categorized as “early” and “middle” studies, respectively. A study analyzed VE during 01 Jul 2021 to 31 Oct 2021 was categorized as “Late study”. Severe COVID-19 was defined as hospital admission with the exception of the *Chon et al study which defined severe COVID-19 as death. Circle symbols represents data for young adults (<65 years of age) and triangle symbols represents data for older adults (≥65 years of age). VE: vaccine effectiveness, CI: confidence interval.

### VE among Young Adults

Two studies examined VE against SARS-CoV-2 infection among young adults (Table 1) ^8, 9^. Estimate of VE against SARS-CoV-2 infection was 89% in an early study ^8^, and 73% in a middle period study ^9^. Five studies examined mRNA VE against severe COVID-19 among young adults (Table 1) ^9, 11-14^. Estimate of VE against COVID-19 hospitalization ranged from 75–97% and 91– 95% among an early study and middle studies, respectively ^9-13^. A late study evaluated VE against severe COVID-19 defined as death and reported VE for young adults that ranged from 81.5% (for mRNA1273) to 84.3% (for BNT162b2) (Figure 2) ^14^.

## Discussion

To our knowledge, ours is the first review to report age-disaggregated VE of SARS-CoV-2 mRNA vaccines focusing on durability of VE in older adult. We found considerable declines in VE against SARS-CoV-2 infection in older adults (≥65 years of age) among the middle studies, which evaluated VE up to 08 August 2021. Older adults also demonstrated decline in VE against severe COVID-19 during this period. Furthermore, marked declines in VE against severe COVID-19 were reported in a late study among a cohort of veterans ≥65 years of age. The study demonstrated that VE against death dropped to 70% for BNT162b2 and 76% for mRNA1273 ^14^. These data contribute to the evidence base for the recent FDA EUA and CDC recommendation of a booster dose of COVID-19 vaccine in older adults ≥65 years of age, and should be considered in the context of significantly higher risk of severe illness from COVID-19 for the older population.

It is important to focus on VE against severe COVID-19 in older adults. Increasing age is the most attributable risk factor of COVID-19-related death among non-vaccinated population ^3^. Furthermore, several articles have reported weaker vaccine-induced immune responses in older adults, including lower concentrations of neutralizing antibodies, compared to younger adults ^16-20^. Although mRNA vaccines were highly effective against severe COVID-19 in older adults after the immunization, the current study suggests that waning VE might lead to severe outcome including death.

We also captured VE of younger adults from eligible studies. We found that the VE against severe COVID-19 maintained >90% in young adults while older adults showed substantial decline resulted in ≤90% of VE in the middle studies. Although these data suggest that VE may wane in older adults faster than young adults, we should be cautious when interpreting the current data because the observation periods after vaccination might differ between young and older adults. For example, the SARS-CoV-2 vaccine campaign in Massachusetts prioritized older adults, starting their immunization in 01 February 2021 whereas those <55 years of age began receiving SARS-CoV-2 vaccines in 19 April 2021 ^21^. Because we did not perform a formal meta-analysis within this paper, we do not attempt to conclude whether VE is waning significantly in older adults compared to younger adults. Furthermore, a study included in our review, which analyzed VE between July to October 2021 showed waning VE against death in veterans <65 years to 81.5-84.3% ^14^. We should bear in mind that the veterans analyzed in the study had a skewed distribution of sex and age. Nonetheless, these results suggest that VE against severe COVID-19 and death is also waning in younger adults.

There are several limitations to our review. First, since there was little to no information regarding the dominant SARS-CoV-2 strain among the studies, circulating SARS-CoV-2 strains were not accounted for in our review. mRNA vaccine VE against SARS-CoV-2 infection is decreasing with increasing report of breakthrough infections ^4, 5, 14^. This is likely due to both waning immunity occurring months after vaccinations, and an emergence of SARS-CoV-2 variants which become predominant during the study period. Several SARS-CoV-2 variants harboring mutations are associated with decreased mRNA VE against infection and/or severe COVID-19 ^22^. Therefore, changes in circulating SARS-CoV-2 strains might have affected VE in the “middle” and “late” studies. Second, only U.S.-based studies were included. Our review focused on these studies for several reasons: 1) the initiation and progression of vaccination campaigns varied between countries; 2) some countries applied a modified vaccine regimen (e.g. prolonged interval between 1^st^ and 2^nd^ dose); and 3) the predominant circulating SARS-CoV-2 strain differs between countries. Future reviews should broaden scope to include studies conducted outside the U.S. Third, definition of VE (e.g. 7 vs 14 days after second dose, hospitalization vs death) and study design (e.g. test-negative, cohort registry) varied between the studies. Due to the above limitations, we did not pool VE data nor perform meta-analysis to avoid potential bias. Fourth, we only considered VE estimates for young adults when they appeared in the same articles which included estimates of VE in older adults. Finally, some studies had an overlap between their observation periods. We categorized studies as early, middle, and late based on the observation period. However, some of the middle studies had overlapped their study periods with early and late studies.

In summary, we found that VE against SARS-CoV-2 infection and severe COVID-19 wanes in older adults. These results are consistent with recent policies recommending a booster dose of COVID-19 vaccine in older adults ≥65 years of age. Overall, our vaccine response to the coronavirus pandemic should consider precision vaccinology principles, such as accounting for age as a key biologic variable.

## Data Availability

All data produced in the present work are contained in the manuscript.

## Acknowledgments

We thank our fellow faculty within the *Precision Vaccines Program* (PVP) for helpful conversations as well as the PVP administrative staff for their support. The PVP is supported in part via NIH/NIAID grants and contracts including Adjuvant Discovery (HHSN272201400052C and 75N93019C00044) and Development (HHSN272201800047C) Program Contracts as well as the BCH Department of Pediatrics.

## Declaration of Interest Statement

OL is a part time employee of the U.S. FDA and a named inventor on patents relating to vaccine adjuvants and human *in vitro* models that model vaccine action.

## Notes

### Funding Statement

This study is funded by in part via NIH/NIAID NIH/NIAID grants and contracts including Adjuvant Discovery (HHSN272201400052C and 75N93019C00044) and Development (HHSN272201800047C) Program Contracts as well as the Boston Children's Hospital Department of Pediatrics.

### Author Declarations

This study reviewed 8 peer-reviewed publication available in PubMed. Descriptions of the included papers are stated in the paper.

